# Time between Symptom Onset, Hospitalisation and Recovery or Death: a Statistical Analysis of Different Time-Delay Distributions in Belgian COVID-19 Patients

**DOI:** 10.1101/2020.07.18.20156307

**Authors:** C. Faes, S. Abrams, D. Van Beckhoven, G. Meyfroidt, E. Vlieghe, N. Hens, On behalf of The Belgian Collaborative Group on COVID-19 Hospital Surveillance

**Author notes:** Members of the Belgian Collaborative Group on COVID-19 Hospital Surveillance: Amir-Samy Aouachria, Kristof Bafort, Leïla Belkhir, Nathalie Bossuyt, Vincent Colombie, Nicolas Dauby, Paul De Munter, Jessika Deblonde, Didier Delmarcelle, Mélanie Delvallee, Rémy Demeester, Thierry Dugernier, Xavier Holemans, Benjamin Kerzmann, Pierre Yves Machurot, Philippe Minette, Jean-Marc Minon, Saphia Mokrane, Catherine Nachtergal, Séverine Noirhomme, Denis Piérard, Camelia Rossi, Carole Schirvel, Erica Sermijn, Frank Staelens, Filip Triest, Nina Van Goethem, Jens Van Praet, Anke Vanhoenacker, Sarah Cooreman, Elise Willems, Chloé Wyndham-Thomas.

## Abstract

**Background:** There are different patterns in the COVID-19 outbreak in the general population and amongst nursing home patients. Different age-groups are also impacted differently. However, it remains unclear whether the time from symptom onset to diagnosis and hospitalization or the length of stay in the hospital is different for different age groups, gender, residence place or whether it is time dependent.

**Methods:** Sciensano, the Belgian Scientific Institute of Public Health, collected information on hospitalized patients with COVID-19 hospital admissions from 114 participating hospitals in Belgium. Between March 14, 2020 and June 12, 2020, a total of 14,618 COVID-19 patients were registered. The time of symptom onset, time of COVID-19 diagnosis, time of hospitalization, time of recovery or death, and length of stay in intensive care are recorded. The distributions of these different event times for different age groups are estimated accounting for interval censoring and right truncation in the observed data.

**Results:** The truncated and interval-censored Weibull regression model is the best model for the time between symptom onset and diagnosis/hospitalization best, whereas the length of stay in hospital is best described by a truncated and interval-censored lognormal regression model.

**Conclusions:** The time between symptom onset and hospitalization and between symptom onset and diagnosis are very similar, with median length between symptom onset and hospitalization ranging between 3 and 10.4 days, depending on the age of the patient and whether or not the patient lives in a nursing home. Patients coming from a nursing home facility have a slightly prolonged time between symptom onset and hospitalization (i.e., 2 days). The longest delay time is observed in the age group 20-60 years old. The time from symptom onset to diagnosis follows the same trend, but on average is one day longer as compared to the time to hospitalization. The median length of stay in hospital varies between 3 and 10.4 days, with the length of stay increasing with age. However, a difference is observed between patients that recover and patients that die. While the hospital length of stay for patients that recover increases with age, we observe the longest time between hospitalization and death in the age group 20-60. And, while the hospital length of stay for patients that recover is shorter for patients living in a nursing home, the time from hospitalization to death is longer for these patients. But, over the course of the first wave, the length of stay has decreased, with a decrease in median length of stay of around 2 days.

## 1 Introduction

The world is currently faced with an ongoing coronavirus disease 2019 (COVID-19) pandemic. The disease is caused by the severe acute respiratory syndrome coronavirus 2 (SARS-CoV-2), a new strain of the coronavirus, which was never detected before in humans, and is a highly contagious infectious disease. The first outbreak of COVID-19 occurred in Wuhan, province Hubei, China in December 2019. Since then, several outbreaks have been observed throughout the world. On February 21, 2020, a cluster of COVID-19 cases was confirmed in Italy, the first European country affected by the virus. One week later, several imported cases were reported in Belgium, after a week of school holidays. As from March 7, the first generation of infected individuals as a result of local transmission was confirmed in Belgium.

There is currently little detailed knowledge on the time interval between symptom onset and hospital admission, nor on the length of stay in hospital. However, information about the length of stay in hospital is important to predict the number of required hospital beds, both for beds in general hospital and beds in the intensive care unit (ICU), and to track the burden on hospitals (Vekaria et al. 2020). The time delay from illness onset to death is important for the estimation of the case fatality ratio (Donnely et al., 2003). Individual-specific characteristics, such as, e.g., the gender, age and co-morbidity of the individual, could potentially explain differences in length of stay in the hospital and are therefore important to correct for.

Therefore, in the present study, we investigate the time of symptom onset to hospitalization and the time of symptom onset to diagnosis, as well as the length of stay in hospital. More specifically, we consider and compare parametric distributions for these event times enabling to appropriately take care of truncation and interval censoring. In Section 2, we introduce the epidemiological data and the statistical methodology used for the estimation of the parameters associated with the aforementioned delay distributions. The results are presented in Section 3 and avenues of further research are discussed in Section 4.

## 2 Methods

### 2.1 Clinical surveillance of COVID-19 hospitalized patients

The hospitalized patients clinical database is an ongoing multicenter registry that collects information on hospital admission related to COVID-19 infection. The data are regularly updated as more information from the hospitals are sent in. At the time of writing this manuscript, the data were available until June 12, 2020. The individual patients’ data are collected through 2 online questionnaires: one with data on admission and one with data on discharge. Data are reported for all hospitalized patients with a confirmed COVID-19 infection. The reporting is strongly recommended by the Belgian Risk Management Group, therefore the reporting coverage is high (> 70% of all hospitalized COVID-19 cases) (Van Goethem et al. 2020).

In the survey, there is information about 14,618 patients, hospitalized between March 1, 2020 and June 12, 2020, including age and gender. From these, 6,866 of the hospitalized patients are females and 7,752 are males. 258 hospitalized patients are less than 20 years old, 4,338 individuals are between 20 and 59 years of age, 5,480 are between 60 and 79 years of age and 4,542 have an age above 80 years. From these patients, it is known that 2,337 live in a nursing home and 6,812 do not. Table 1 shows that a large proportion of the hospitalized 60+ patients are known to live in a nursing home facility (about 12% for patients aged 60-79 and 35% for patients aged 80+). As expected, below the age of 60 years, there is only a very small proportion of patients that come from a nursing home facility.

**Table 1:**
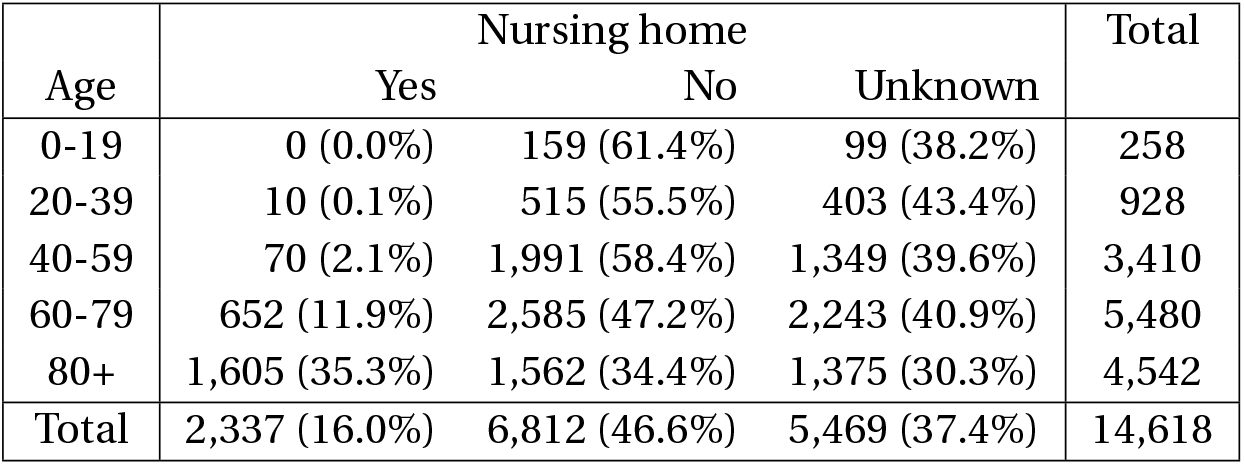
Number of hospitalized patients in the national surveillance survey of COVID-19 hospitalizations, between March 1, 2020 and June 12, 2020.

The survey contains information on 1,831 patients hospitalized during the initial phase of the outbreak (between March 1 and March 20); 4,998 patients in the increasing phase of the outbreak (between March 21 and March 31); 5,094 in the descending phase (between April 1 and April 18); and 2,695 individuals at the end of the first wave of the COVID-19 epidemic (between April 19 and June 12). The time trend in the number of hospitalizations is presented in Figure 1. Black dots represent the number of patients included in the national surveillance survey and the red dots show the reported number of confirmed hospitalizations in the population. The time trend in the survey matches well with the time trend of the outbreak in the whole population, though with some under-reporting in April and May.

**Figure 1:**
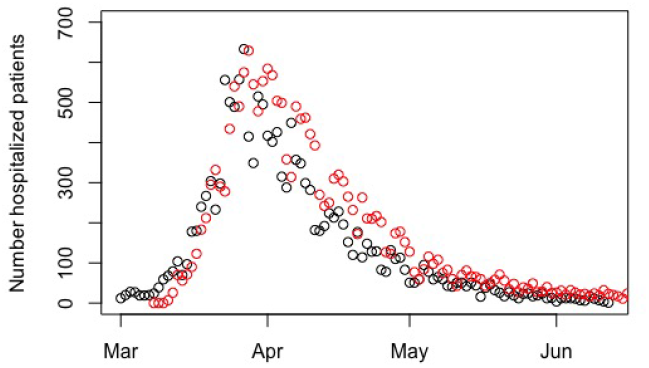
Observed number of new hospitalization in the national surveillance survey (black) and reported in the whole population (red)

The date variables were checked for consistency. Observations identified as inconsistent were excluded for analyses related to the inconsistent dates. A flow diagram of the exclusion criteria is displayed in Figure 2. The time of symptom onset and time of hospitalization is available for 13,321 patients. The date of symptom onset is determined based on the patient anamnesis history made by the clinicians. Patients that were hospitalized before the start of symptoms (i.e., 715 patients) were not included. These include patients with nosocomial infections admitted prior to COVID-19 infection for other long-term pathologies, then got infected at hospital and developing COVID-19-related symptoms long after admission. Patients reporting a delay between symptoms and hospitalization of more than 31 days (i.e., 121 patients) were also not included, because it is unclear for these patients whether the reason for hospital admission was COVID-19 infection. A sensitivity analysis including patients with event times above 31 days is conducted. Patients with missing information on age (i.e., 12 patients) or gender (i.e., 109 patients) were not included in the statistical analysis. This resulted in a total of 12,364 patients which were used to estimate the distribution of the time between symptom onset and hospitalization.

**Figure 2:**
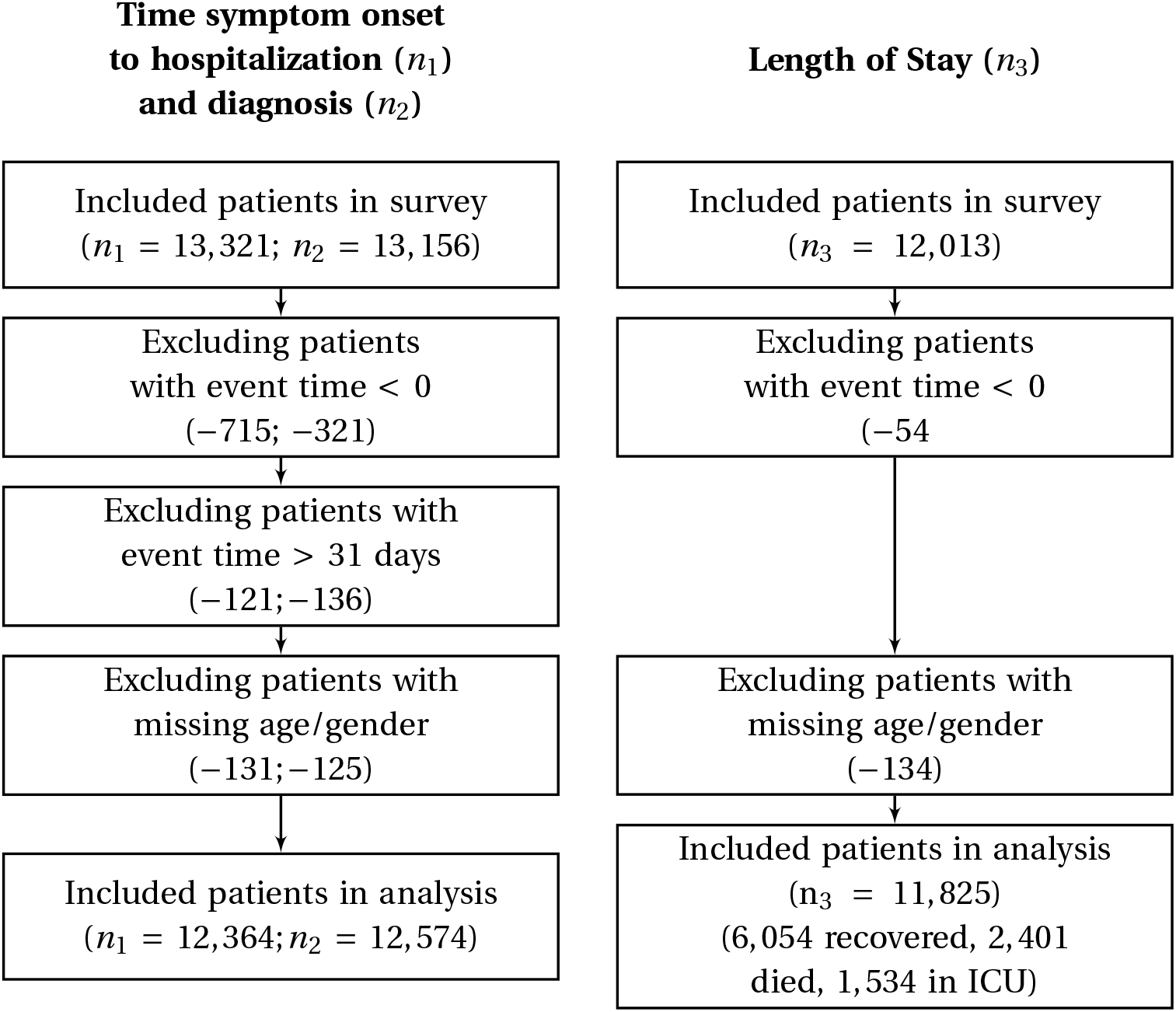
Flow diagram: (*n*_1_; *n*_2_) on the left hand side correspond, respectively, to the number of patients for the analysis of time of symptom onset to hospitalization and for time of symptom onset to diagnosis. The number of patients *n*_3_ on the right hand size correspond to the length of stay in hospital.

The time of symptom onset and time of diagnosis is available for 13,156 patients. Some of these were diagnosed prior to having symptoms (321) or experienced symptoms more than 31 days before diagnosis (136), and are excluded as these might be errors in reporting dates. Similarly, the delay between symptoms and detection time is truncated at 31 days; but a sensitivity analysis including these patients is performed. In total, 125 patients were removed because of missing information on age and/or gender, resulting in 12,574 patients used in the analysis of the time from symptom onset to diagnosis.

The time between hospitalization and discharge from hospital is available for 12,013 patients, either discharged alive or dead. For patients that were hospitalized before the start of symptoms (i.e., 528 patients), we use the time between the start of symptoms and discharge. Patients with negative time intervals (54 patients) are excluded for further analysis. Another 134 patients were discarded because of missing covariate information with regard to their age or gender. From these patients, we know that 6,054 recovered from COVID-19, while 2,401 died. From the hospitalized patients, there is information about the length of stay at ICU for 1,534 patients.

Note that we analyzed an anonymized subset of data from the hospital COVID-19 clinical surveillance database of the Belgian public health institute Sciensano. Data from Sciensano was shared with the first author through a secured data transfer platform.

### 2.2 Descriptive analysis of the delay times

As there exist large differences between healthcare systems in different countries, the reporting lag and time to hospitalization can be very different amongst countries (WHO, 2020). In this section, we describe the observed delay from symptom onset to hospitalization, from symptom onset to diagnosis and the length of stay in hospital during the first wave of COVID-19 infections in Belgium. Statistical analysis results thereof are presented in Section 3.

The observed distribution of the delay from symptom onset to hospitalization (left panel) and to diagnosis (right panel) is presented in Figure 3. The observed length of stay in hospital and in ICU is presented in Figure 4 for all patients as well as separately for those that died or recovered from the disease.

**Figure 3:**
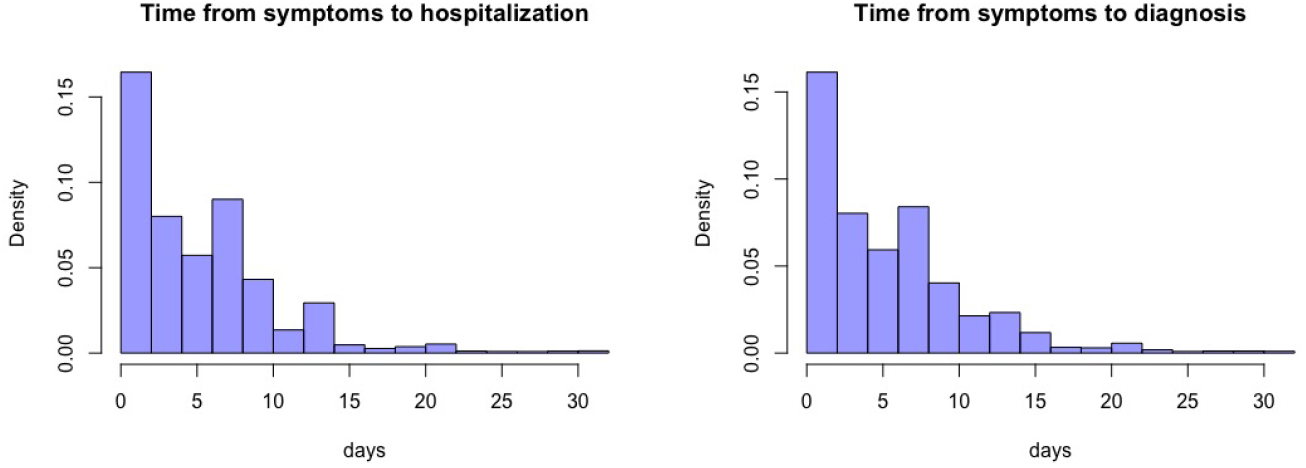
Observed density (proportion of the population in the survey) of time between symptom onset and hospitalization (left) and between symptom onset and diagnosis (right)

**Figure 4:**
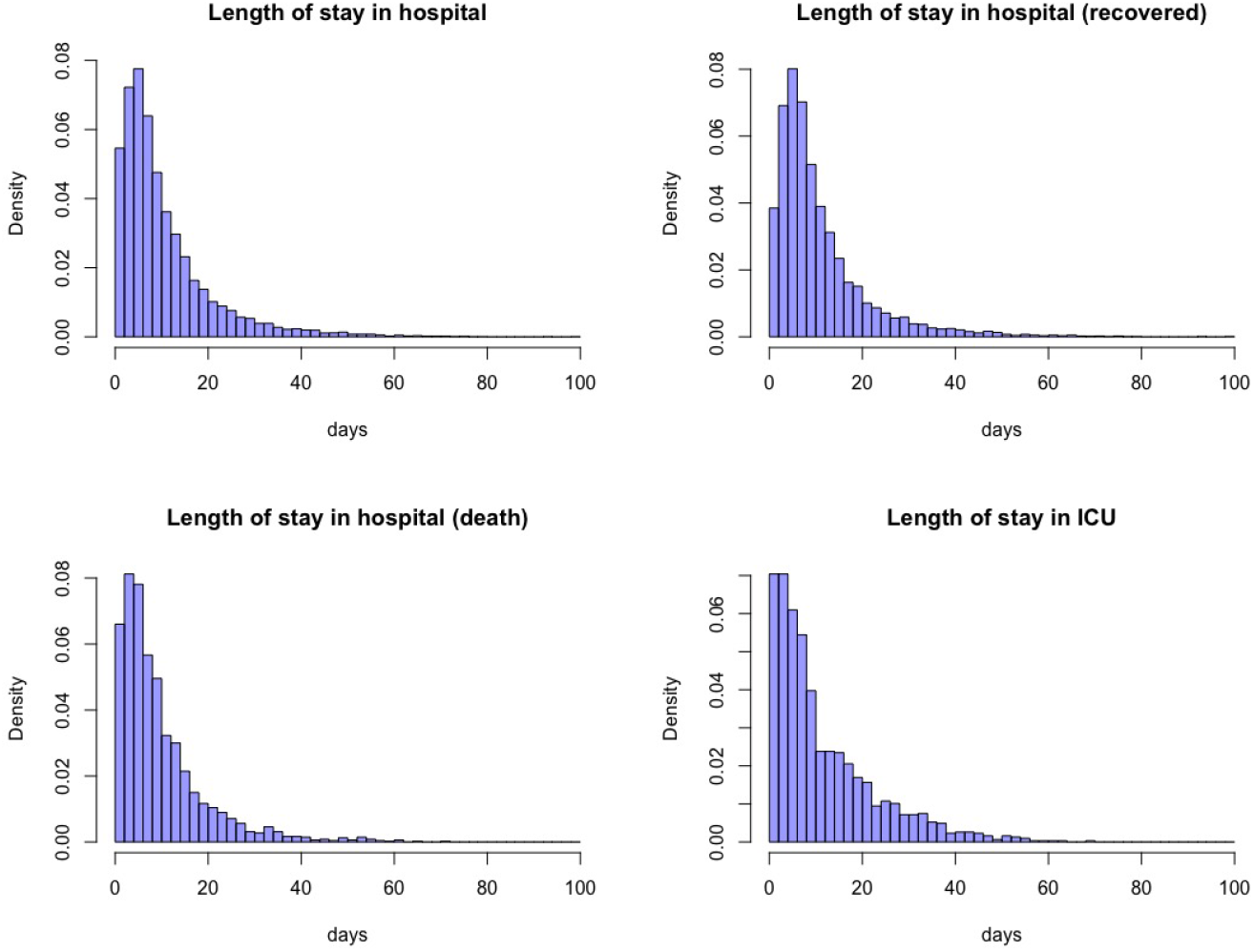
Observed length of stay in hospital (all COVID-19 patients (top left), recovered patients (top right), patients that died (bottom left) and length of stay in ICU (bottom right)

Summary information about these distributions is presented in Tables A1 and A2 in the Appendix. Note that the empirical distributions shown in in Figure 4 do not explicitly account for truncation of the event times at the end of the study. More specifically, the relative frequencies of short-term stays in the hospital are inflated by the absence of patients with larger lengths of stay that are still in hospital at the end of the study period, and therefore missing in the data. Consequently, these graphs should be interpreted with care.

While the observed delay between symptom onset and hospitalization is between 0 and 31 days, 75% of the hospitalizations occur within 8 days after symptom onset. This is however shorter in the youngest age group (<20 years) and in the elderly group (>90 years). Also patients coming from nursing homes seem to be hospitalized faster as compared to the general population. Over the course of the first wave, the observed time between symptom onset and hospitalization was largest in the increasing phase of the epidemic (between March 21 and March 30). The time between symptom onset and diagnosis is very similar, ranging between 0 and 31 days, with 75% of the diagnoses occurring within 8 days after symptom onset. It should be noted that these observations are based on hospitalized patients, and non-hospitalized patients might have a quite different evolution in terms of their symptoms. As non-hospitalized patients were rarely tested in the initial phase of the epidemic, no conclusions can be made for this group of patients.

The observed median length of stay in hospital is 8 days, with 95% of the patients have values ranging between 1 and 40 days. 25% of the patients stay longer than 14 days in the hospital. The median length of stay seems to increase with age (from 3 days in age group < 20 to 6 in age group 20− 80, 9 in age group 80− 90 and 10 days in age group > 90). On the other hand, with time since introduction of the disease in the population, the length of stay seems to decrease, though this might be biased due to incomplete reporting of LOS in patients who are actually still admitted at the time of writing. Therefore, these observed statistics should be interpreted with care. Similar results are observed for the length of stay in ICU.

### 2.3 Statistical Model

Different flexible parametric non-negative distributions can be used to describe the delay distributions, such as the exponential, Weibull, lognormal and gamma distributions (Held et al., 2020). However, as the reported event times are expressed in days, the discrete nature of the data should be accounted for in the estimation of the distributional parameters with regard to the respective delay distributions. Different techniques are used in literature to take this into account. Donnely et al. (2003) and Hens et al. (2012) assume a discrete probability distribution parameterized by a continuous distribution. Alternatively, Cowling et al. (2009) estimate the serial interval using interval censoring techniques from survival analysis. Reich et al. (2009) and Linton et al. (2020) use doubly interval-censoring methods for estimation of the incubation distribution. We use interval-censoring methods originating from survival analysis to deal with the discrete nature of the data, to acknowledge that the observed time is not the exact event time (Sun, 2006). Let *x*_*i*_ be the recorded event time. Instead of assuming that *x*_*i*_ is observed exactly, it is assumed that the delay is in the interval (*L*_*i*_, *R*_*i*_), with *L*_*i*_ = *x*_*i*_ − 0.5 and *R*_*i*_ = *x*_*i*_ +0.5 for *x*_*i*_ ≥ 1 and *L*_*i*_ = = 10^− 3^ and *R*_*i*_ = 0.5 for *x*_*i*_ = 0. As a sensitivity analysis, we compare this assumption with the wider interval (*L*_*i*_ = *x*_*i*_ − 1, *R*_*i*_ = *x*_*i*_ + 1).

In addition, the delay distribution is often truncated, either because there is a maximal clinical delay period (e.g., time between symptom onset and hospitalization is at most 31 days) or because the hospitalization is close to the end of the study (e.g., if hospitalization is 14 days before the end of the study, the observed length of stay cannot exceed 14 days) and partial information about patients still being hospitalized is not part of the database. We therefore use a likelihood function accommodating the right-truncated and interval-censored nature of the observed data to estimate the parameters of the distributions (Cowling et al., 2014). The likelihood function is given by

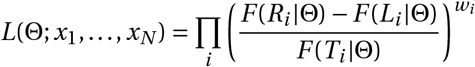

in which *T*_*i*_ is the (individual-specific) truncation time and *F* (·) is the cumulative distribution function corresponding to the density function *f* (·). We truncate the time from symptom onset to diagnosis and the time from symptom onset to hospitalisation to 31 days (*T*_*i*_ ≡ 31). The length of stay in hospital is truncated at *T*_*i*_ = *E* − *t*_*i*_, in which *t*_*i*_ is the time of hospitalization and *E* denoted the end of the study period (June 6, 2020). In addition, to account for possible under-reporting in the survey, each contribution is weighted by the post-stratification weight *w*_*i*_ ≡ *w*_*t*_ defined as 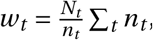, where *t* is the day of hospitalization for patient *i, N*_*t*_ the number of hospitalizations in the population on day *t* and *n*_*t*_ is the number of reported hospitalizations in the survey on day *t*.

We assume a Weibull and lognormal distribution for the delay time distribution. The two parameters of each distribution are regressed on age, gender, nursing home and time period. A maximum likelihood approach is used for parameter estimation. The Bayesian Information Criterion (BIC) is used to select the best fitting parametric distribution and the best regression model among the candidate distributions/models. Only significant parameters are included in the final model. In addition, the delay distributions are summarized by their estimated mean, median, 5th, 25th, 75th and 95th quantiles, as these can be helpful in guiding policy decision making and future COVID-19 modeling approaches.

## 3 Results

### 3.1 Symptom Onset to Hospitalization and to Diagnosis

Overall, the delay between symptom onset and hospitalization can be described by a truncated Weibull distribution with shape parameter 0.845 and scale parameter 5.506. The overall average delay is very similar to the one obtained by Abrams et al. (2020), based on a stochastic discrete time model relying on an Erlang delay distribution. However, there are significant differences in the time between symptom onset and hospitalization between males and females, among different age groups, between living statuses (nursing home, general population or unknown) and between different reporting periods. As the truncated Weibull distribution has a lower BIC as compared to the lognormal distribution (BIC of 66,923 and 68,657 for Weibull and lognormal distributions, respectively), results for the Weibull distribution are presented. In Table 2, the regression coefficients of the scale (*λ*) and shape parameters (*γ*) of the Weibull distribution are presented. The impact on the time between symptom onset and hospitalization is visualized in Figure 5, showing the model-based 5%, 25%, 50%, 75% and 95% quantiles of the delay times.

**Table 2:** Summary of the regression of the scale (*λ*) and shape (*γ*) parameters for reported delay time between symptom onset and hospitalization and between symptom onset and diagnosis, based on a truncated Weibull distribution: parameter estimate, standard error and significance (^∗^ corresponds to p-value< 0.05; ^∗∗^ to p-value < 0.01 and ^∗∗∗^ to < 0.001). The reference group used are females of age > 80 living in nursing home that are hospitalized in the period 01-03 to 20-03.

| | $\log(\lambda)$ | | $1/\gamma$ | |
| --- | --- | --- | --- | --- |
|  | Est | s.e. | Est | s.e. |
| Symptom onset - hospitalization |  |  |  |  |
| (Intercept) | 1.218 | 0.044*** | 1.208 | 0.036*** |
| Male | 0.032 | 0.020 | -0.076 | 0.017*** |
| Age < 20 | -0.535 | 0.104*** | 0.033 | 0.084 |
| Age 20 – 60 | 0.618 | 0.034*** | -0.490 | 0.028*** |
| Age 60 – 80 | 0.433 | 0.035*** | -0.251 | 0.029*** |
| Nursing home: No | -0.452 | 0.044*** | 0.250 | 0.039*** |
| Nursing home: Unknown | -0.046 | 0.021* | 0.143 | 0.019*** |
| Period 21-03 to 30-03 | 0.234 | 0.033*** | -0.100 | 0.028*** |
| Period 31-03 to 18-04 | 0.187 | 0.035*** | 0.137 | 0.030*** |
| Period 19-04 to 12-06 | 0.028 | 0.047 | 0.495 | 0.039*** |
| Symptom onset - diagnosis |  |  |  |  |
| (Intercept) | 1.527 | 0.042*** | 1.232 | 0.035*** |
| Male | 0.040 | 0.019* | -0.098 | 0.016*** |
| Age < 20 | -0.400 | 0.081*** | -0.196 | 0.059*** |
| Age 20 – 60 | 0.383 | 0.031*** | -0.389 | 0.025*** |
| Age 60 – 80 | 0.284 | 0.031*** | -0.201 | 0.026*** |
| Nursing home: No | -0.480 | 0.038*** | 0.062 | 0.031* |
| Nursing home: Unknown | 0.022 | 0.019 | -0.004 | 0.016 |
| Period 21-03 to 30-03 | 0.181 | 0.034*** | -0.187 | 0.030*** |
| Period 31-03 to 18-04 | 0.029 | 0.036 | 0.097 | 0.032** |
| Period 19-04 to 12-06 | -0.297 | 0.047*** | 0.545 | 0.042*** |

**Figure 5:**
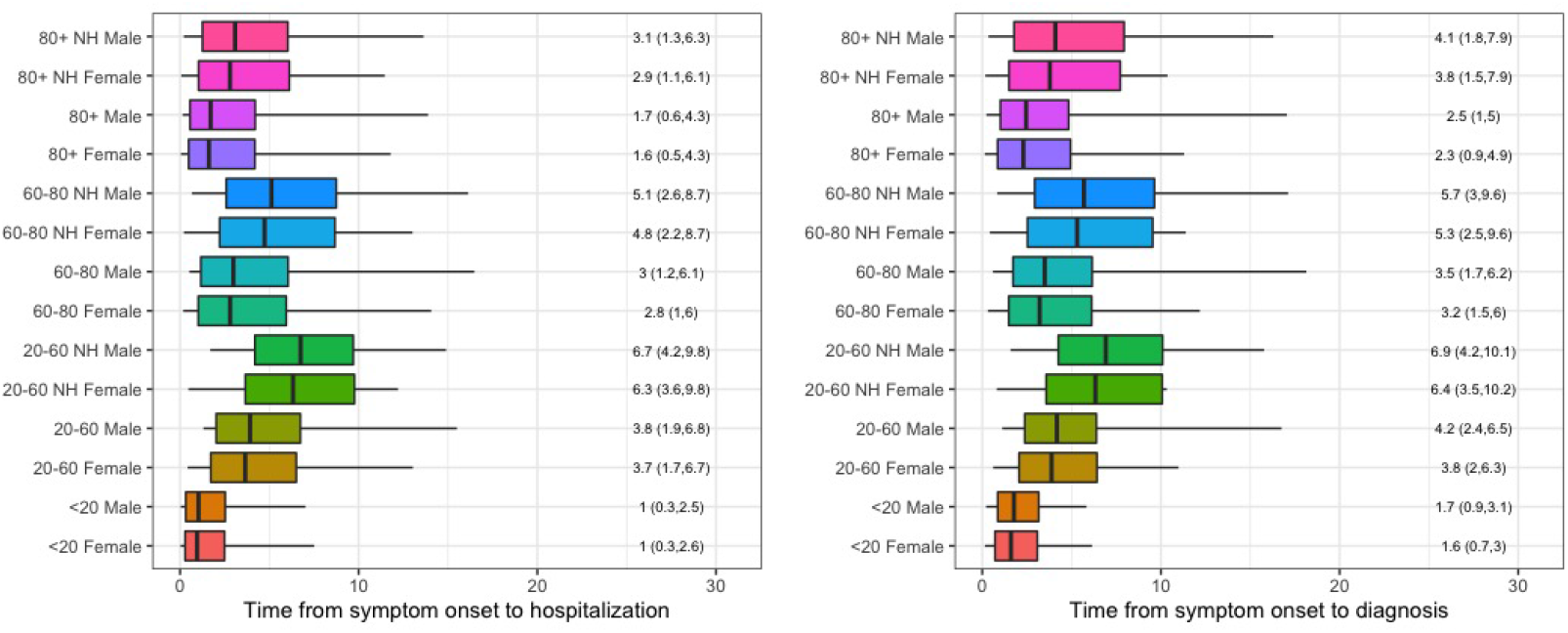
Comparison of delay distribution for different population groups. The boxplots show the estimated 5%, 25%, 50%, 75% and 95% quantiles, based on Weibull regression. Results are based on period 21-03 to 30-03. The reported values correspond to the 50%(25%, 75%) quantiles.

Age has a major impact on the delay between symptom onset and hospitalization, with the youngest age group having the shortest delay (median of 1 day, but with a quarter of the patients having a delay longer than 2.6 days). The time from symptom onset to hospitalization is more than doubled in the age groups 20-60 and 60-80 as compared to the age group < 20 (median close to 4 days and a delay of more than 6.7 days for a quarter of the patients). In contrast the increase in time between symptom onset and hospitalization is 50% in the age group 80+ as compared to the youngest age group < 20 (median delay of 1.6 days, with a quarter of the patients having a delay longer than 4.3 days). After correcting for age, it is observed that the time delay is somewhat higher when patients come from a nursing home facility, with an increase of approximately 2 days. Note that in the descriptive statistics, we observed shorter delay times for patients coming from nursing homes. This stems from the fact that 80+ year old’s have shorter delay times as compared to patients of age 20-79, but the population size in the 80+ group is much larger as compared to the 20-79 group in nursing homes. This is known as Simpson’s paradox. And although statistical significant differences were found for gender and period, we observe very similar time delays between males and females and in the different time periods (see Figure A4 in the Appendix). Note, however, that there are indeed differences, but mainly in the tails of the distribution; with, e.g., the 5% longest delay times between symptoms and hospitalizations observed for males.

The time between symptom onset and diagnosis is also best described by a truncated Weibull distribution (shape parameter 0.900, scale parameter 5.657). As again the truncated Weibull distribution has a lower BIC value as compared to the lognormal distribution (BIC values of 68,106 and 69,652 for Weibull and lognormal, respectively), results for the Weibull distribution are presented. Parameter estimates are very similar to the distribution for symptom onset and hospitalization, and are presented in Table 2. The median delay between symptom onset and diagnosis is approximately one day longer as compared to the median delay between symptom onset and hospitalization. The diagnosis was typically made upon hospital admission to confirm COVID-19 infection. This is why the date of admission is very close to date of diagnosis. The same effects of age and nursing home are found for the time between symptom onset and diagnosis, as compared to the time to hospitalization. Especially at the increasing phase of the epidemic, the time between symptom onset and diagnosis was longer as compared to the time between symptom onset and hospitalization (see Figure A4), but this delay has shortened over time.

As a sensitivity analysis, a comparison is made with an analysis without truncating the time between symptom onset and hospitalisation or diagnosis. Results are presented in Figure A5, and are very similar to the once presented here. In addition, a sensitivity analysis assuming that the time delay is interval censored with time intervals defined as (*x*_*i*_ − 1, *x*_*i*_ +1) is presented in Figure A3, yielding very similar results. It was also investigated whether or not there a difference between neonati (with virtually no symptoms, but diagnosed at the time of birth or at the time of the mothers testing prior to labour) and other children. For all children < 20 years of age, we found a median time from symptom onset to hospitalization and diagnosis to be 1 and 1.6 days, respectively. If we only consider children > 0 years of age, a small increase is found (1.5 (0.5-3.4) days for time to hospitalization and 1.8 (0.7-3.7) for time to diagnosis).

### 3.2 Length of Stay in Hospital

A summary of the estimated length of stay in hospital and ICU is presented in Table 3 and Figure 6 based on the lognormal distribution. The lognormal distribution has a slightly smaller BIC value as compared to the Weibull distribution for the length of stay in hospital (BIC value of 76,928 for Weibull and 76,865 for lognormal) and for the length of stay in ICU (BIC value of 7,341 for Weibull and 7,312 for lognormal).

**Table 3:** Summary of the regression of the log-mean (*µ*) and log-standard deviation (*σ*) parameters for the length of stay in hospital and ICU, based on the lognormal distribution: parameter estimate, standard error and significance (^∗^ corresponds to p-value< 0.05; ^∗∗^ to p-value < 0.01 and ^∗∗∗^ to < 0.001). The reference group used are females of age > 80 living in nursing home that are hospitalized in the period 01-03 to 20-03.

| | $\mu$ | | $\sigma$ | |
| --- | --- | --- | --- | --- |
|  | Est | s.e. | Est | s.e. |
| Length of stay in hospital |  |  |  |  |
| (Intercept) | 2.480 | 0.035*** | 0.913 | 0.024*** |
| Male | 0.091 | 0.019*** | / | / |
| Age < 20 | -1.143 | 0.067*** | / | / |
| Age 20 – 60 | -0.553 | 0.024*** | / | / |
| Age 60 – 80 | -0.087 | 0.023*** | / | / |
| Period 21-03 to 30-03 | -0.242 | 0.034*** | -0.008 | 0.027 |
| Period 31-03 to 18-04 | -0.270 | 0.034*** | 0.017 | 0.027 |
| Period 19-04 to 12-06 | -0.359 | 0.038*** | 0.087 | 0.030** |
| Length of stay in ICU |  |  |  |  |
| (Intercept) | 2.183 | 0.152*** | 1.052 | 0.029*** |
| Age < 20 | -0.443 | 0.401 | / | / |
| Age 20 – 60 | 0.098 | 0.126 | / | / |
| Age 60 – 80 | 0.273 | 0.122* | / | / |
| Nursing home: No | -0.282 | 0.147 | / | / |
| Nursing home: Unknown | -0.170 | 0.072* | / | / |
| Period 21-03 to 30-03 | -0.135 | 0.112 | / | / |
| Period 31-03 to 18-04 | -0.186 | 0.115 | / | / |
| Period 19-04 to 12-06 | -0.418 | 0.151** | / | / |

**Figure 6:**
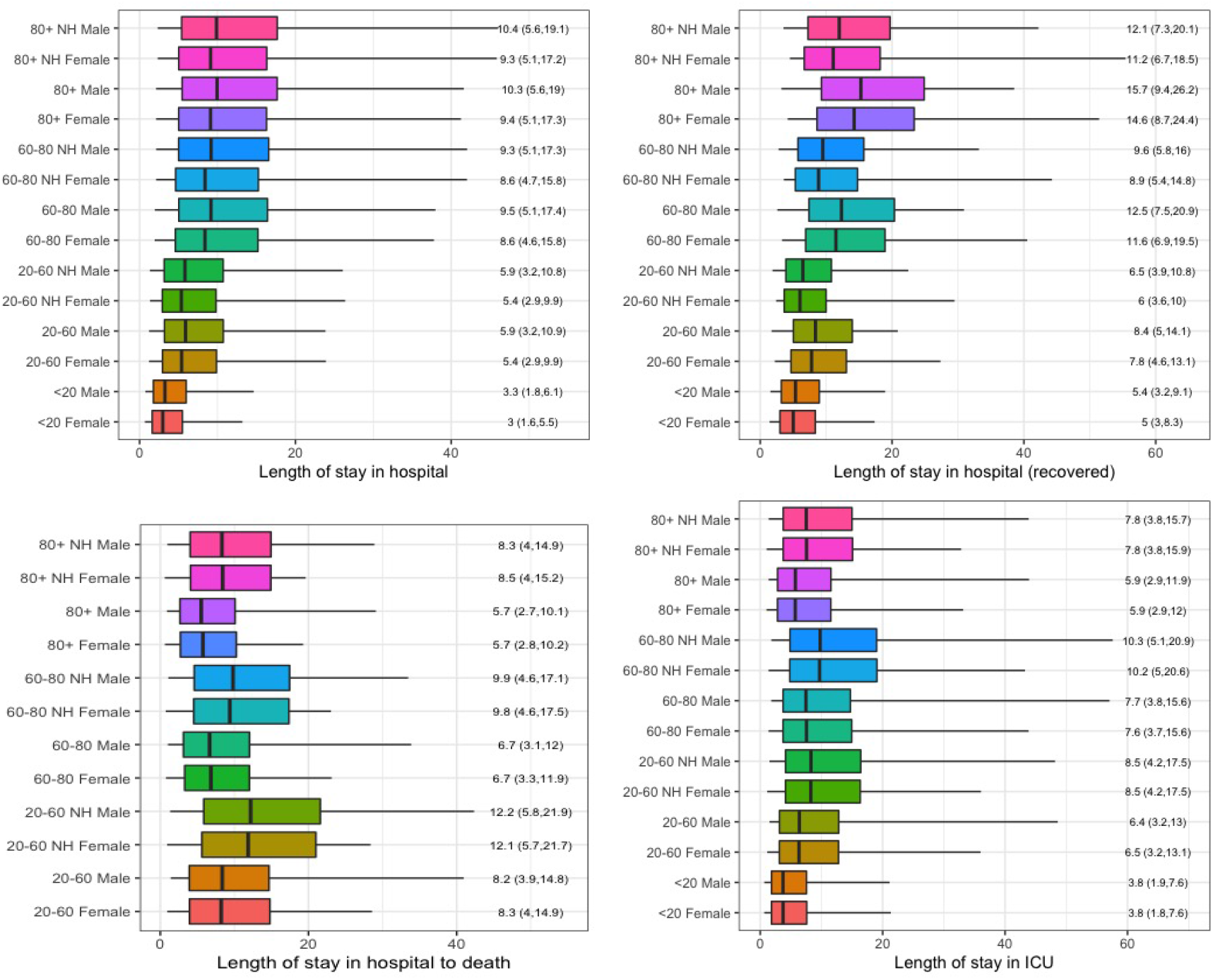
Comparison of hospital length of stay for different groups. Top left is LOS overall, top right is LOS for recovered patients, bottom left is LOS for patients that died and bottom right is LOS in ICU. The boxplots show the 5%, 25%, 50%, 75% and 95% quantiles.

The median length of stay in hospital is close to 3 days in the age group less than 20 years old, but 25% of these patients stay longer than 5.5 days in hospital for females and more than 8.6 days for males and 5% thereof stay longer than 13 days for females and 14 days for males. The length of stay increases with age, with a median length of stay of around 5.4 days for females aged 20-60 and 5.9 days for males aged 20-60. A quarter of the patients in age group 20-60 stay longer than 10 days and 5% stay longer than 24 days. This further increases for patients above 60 years of age, with a median length of stay of around 8.6 and 9.4 days for female and male patients aged 60-80 years and 9.4 and 10.3 days for female and male patients above 80 years of age. A large proportion of the elderly patients stay much longer in hospital. A quarter of these patients stay longer than 15.7-17.4 days for patients of age 60-80 years and longer than 17.3-19 days for patients of age above 80. Some very long hospital stays are observed in this age group, with 5% of the stays being longer than 38 and 41 days for females and males in the age group 60-80 years, and 42 and 46 days in the age group 80+. No significant difference is found for patients coming from nursing homes. Over the course of the first wave, the length of stay has slightly decreased, with a decrease in median length of stay of around 2 days from the first period to later periods.

Note that this result is corrected for possible bias of prolonged lengths of stay being less probable for more recently admitted patients. Therefore, This might be related to improved to better clinical experience and improved treatments.

The length of stay in ICU (based on the lognormal distribution) is on average 3.8 days for patients below 20 years of age, with a quarter of the patients staying longer than 7.6 days in ICU. Similar to length of stay in hospital, also the length of stay in ICU increases with age. The median length of stay in the age group 20-60 years is 6.4, in age group 60-80 7.6, while in age group 80+ it is slightly shorter (5.9 days). Again, it is observed that a quarter of the patients in age group 20-60 stay longer than 13 days in ICU, in age group 60-80 15.6 days and in 80+ 12 days. Patients living in nursing homes stay approximately 2 days longer in intensive care. No major difference is observed in the length of stay in ICU between males and females, though some prolonged stays are observed in males as compared to females. Similar as the overall length of stay in hospital, the length of stay in ICU has decreased over time (with a decrease of 1 days from the first period to the later periods, and an additional 2 days in the last period).

Table 4 summarizes the length of stay in hospital for patients that recovered or passed away. The lognormal distribution has the smallest BIC value for time from hospitalization to recovery and the Weibull distribution for time from hospitalization to death. Figure 6 also summarizes the estimated distributions. For patients that recovered, the length of stay in hospital increased with age (the median age in age group <20 is 5 days, which increases to 8 days in age group 20-60 years, 12 days in age group 60-80 years and 15 days in age group 80+). In contrast to previous results, we observe that patients living in nursing homes leave hospital approximately 1 day faster as compared to the general population. However, the 5% longest stays in hospital before recovery are longer for patients living in nursing homes.

**Table 4:** Summary of the regression of the log-mean (*µ*) and log-standard deviation (*σ*) parameters for length of stay in hospital for recovered patients and patients that died, based on lognormal distribution and weibull distribution: parameter estimate, standard error and significance (^∗^ corresponds to p-value< 0.05; ^∗∗^ to p-value < 0.01 and ^∗∗∗^ to < 0.001). The reference group used are females of age > 80 living in nursing home that are hospitalized in the period 01-03 to 20-03.

| Length of stay in hospital (reason out of hospital = recovered) |  |  |  |  |
| --- | --- | --- | --- | --- |
| | $\mu$ | | $\sigma$ | |
|  | Est | s.e. | Est | s.e. |
| (Intercept) | 2.724 | 0.044*** | 0.720 | 0.024*** |
| Male | 0.077 | 0.022*** | / | / |
| Age < 20 | -1.074 | 0.074*** | / | / |
| Age 20 – 60 | -0.625 | 0.032*** | / | / |
| Age 60 – 80 | -0.231 | 0.031*** | / | / |
| Nursing home: No | 0.261 | 0.038*** | 0.008 | 0.031 |
| Nursing home: Unknown | 0.012 | 0.0245 | 0.099 | 0.018*** |
| Period 21-03 to 30-03 | -0.307 | 0.040*** | 0.034 | 0.027 |
| Period 31-03 to 18-04 | -0.326 | 0.039*** | 0.007 | 0.027 |
| Period 19-04 to 12-06 | -0.388 | 0.045*** | 0.097 | 0.031** |

Table 4: Summary of the regression of the log-mean ( $\mu$ ) and log-standard deviation ( $\sigma$ ) parameters for length of stay in hospital for recovered patients and patients that died, based on lognormal distribution and weibull distribution: parameter estimate, standard error and significance (\* corresponds to p-value < 0.05; \*\* to p-value < 0.01 and \*\*\* to < 0.001).
| Length of stay in hospital (reason out of hospital = death) |  |  |  |  |
| --- | --- | --- | --- | --- |
| | $\log(\lambda)$ | | $1/\gamma$ | |
|  | Est | s.e. | Est | s.e. |
| (Intercept) | 2.573 | 0.070*** | 0.842 | 0.014*** |
| Age 20 – 60 | 0.372 | 0.084*** | / | / |
| Age 60 – 80 | 0.162 | 0.040*** | / | / |
| Nursing home: No | -0.383 | 0.048*** | / | / |
| Nursing home: Unknown | -0.160 | 0.047** | / | / |
| Period 21-03 to 30-03 | -0.142 | 0.069 | / | / |
| Period 31-03 to 18-04 | -0.156 | 0.069* | / | / |
| Period 19-04 to 12-06 | -0.189 | 0.079* | / | / |

But, while the length of stay in hospital for patients that recover increases with age for all age groups, the survival time of hospitalized patients that died is lower for the age groups 60-80 years (median time of 6.7 days) and 80+ (median time of 5.7 days) as compared to the age group 20 − 60 years (median time of 12.1 days). Also large differences are observed amongst patients coming from nursing homes or not, with the time between hospitalization and death being approximately 3 days longer for patients living in a nursing home. No significant differences are found between males and females.

As a sensitivity analysis, a sensitivity analysis assuming that the time delay is interval censored by (*x*_*i*_ − 1, *x*_*i*_ + 1) is presented in Figure A3. Results are almost identical to the previously presented results. It was also investigated whether the smaller duration of hospitalization for < 20 years can be due to the neonati, for which the duration of stay is often determined by duration of postdelivery recovery of the mother. And indeed, the length of stay in hospital for the youngest age group increases slightly if we take out the children of 0 years of age to 4.1(2.2, 7.6) days for males and 3.7(2, 6.9) days for females. The length of stay in hospital for recovered patients increases to 6.4(3.7, 11) days for males and 5.9(3.4, 10.2) days for females of age between 1 and 19 years of age, making it very similar to the 20 − 60 years old patients that recovered. No impact was observed on the length of stay in ICU.

## 4 Discussion

Previous studies in other countries reported a mean time from symptom onset to hospitalization of 2.62 days in Singapore, 4.41 days in Hong Kong and 5.14 days in the UK (Pellis et al., 2020). Other studies report mean values of time to hospitalization ranging from 5 to 9.7 days (Linton et al., 2020, Kraemer et al., 2020 and Ferguson et al., 2020). In Belgium, the mean time from symptom onset to hospitalization overall is 5.74 days, which is slightly longer as compared to the reported delay in other countries, but depending on the patient population, estimates range between 3 and 10.4 days in Belgium. The time from symptom onset to hospitalization is largest in the age group 20-60 years old, followed by the 60-80 years old. If we compare patients within the same age group, it is observed that the time delay is somewhat higher when patients come from nursing home facility, with an increase of approximately 2 days. The time from symptom onset to diagnosis has the same behaviour, with a slightly longer delay as compared to time from symptom onset to hospitalization.

To investigate the length of stay in hospital, we should make a distinction between patients that recover or that die. While the median length of stay for patients that recover varies between 5 days (in the age group < 20) to 15.7 (in the age group 80+), the median length of stay for patients that die varies between 5 days (in the age group 80+) and 12.2 days (in the age group 20 − 60). Over all patients, the median length of stay varies between 3.1 days (in the age group > 20) to 104 (in the age group 80+). In general, it is observed that the length of stay in hospital for patients that recover increases with age, and males need a slightly longer time to recover as compared to females. But, patients living in nursing homes leave hospital sooner as compared to patients in the same age group from the general population. In contrast, the time between hospitalization and death is longest for the age group 20-60 years, with shorter survival time for the age groups 60-80 years and 80+. The length of stay in hospital for patients that die is longer for patients coming form nursing homes, as compared to patients from the same age group from the general population. A similar trend is observed for the length of stay in ICU.

The length of stay in Belgian hospitals is within the range of the once observed in other countries, though especially the length of stay in ICU seems short in Belgian hospitals. Rees et al. (2020) report a median length of stay in hospital of 14 days in China, and of 5 days outside of China. The median length of stay in ICU is 8 days in China and 7 days outside of China (Rees et al. 2020). Vekaria et al. (2020) report estimated length of stay in England for COVID-19 patients not admitted to ICU of 8.4 days and for ICU length of stay of 12.4 days.

Different sensitivity analysis indicated that the results are robust to some of the assumptions made in the modeling. However, alternative methods could still be investigated to improve the estimation of the delay distributions. First, alternative distributions can be used, having more than two parameters and thus more flexibility, e.g., generalized gamma distributions (for which the gamma, exponential and Weibull distributions are special cases). Second, a truncated doubly-interval censored method could be considered to account for the uncertainty in both time points determining the observed delays (and their intervals). Finally, the impact of severity of illness and co-morbidity on the length of stay in hospital is very important. This was not investigated in this study as this information was not made available, but is an important factor to investigate in future analyses.

## Data Availability

Sciensano are the data-owners of the data.

## Appendix

**Figure A1:**
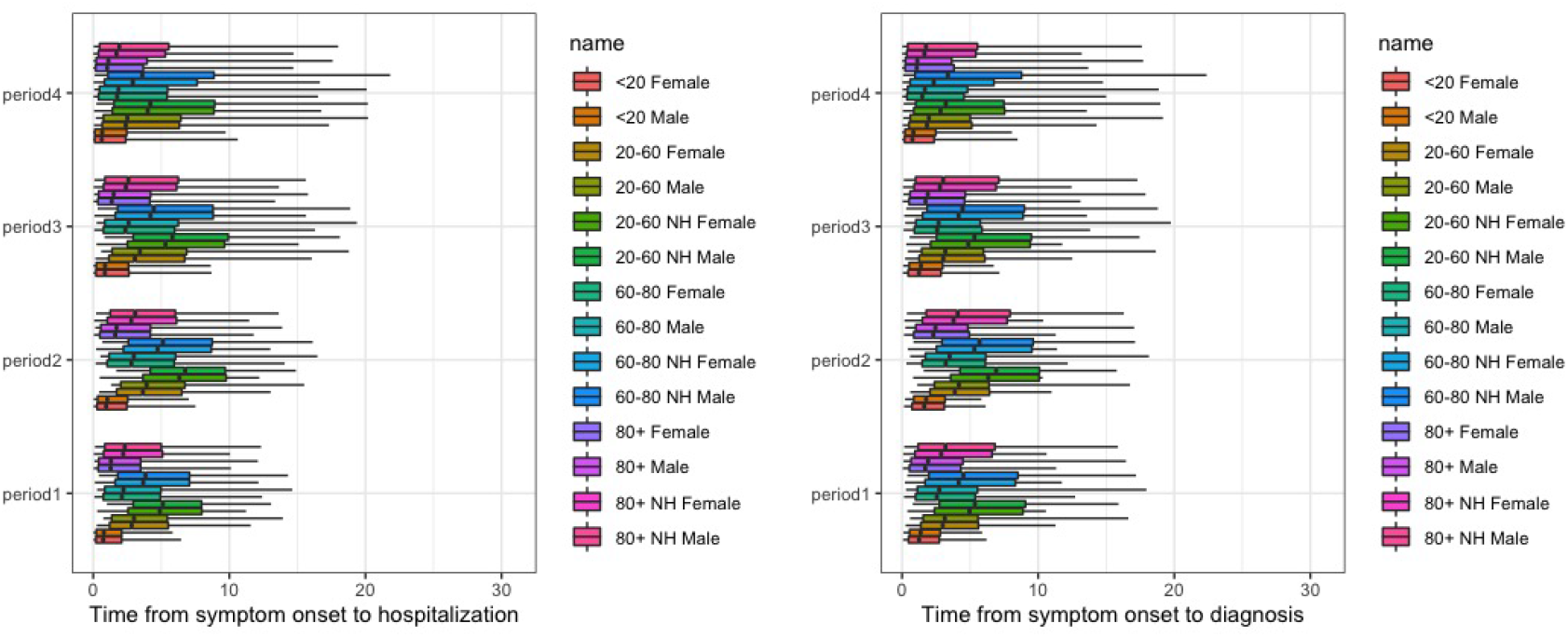
Comparison of delay distribution for different time periods. The boxplots show the estimated 5%, 25%, 50%, 75% and 95% quantiles, based on Weibull regression. Results are based on the time period 01-03 to 20-03 (period 1), 21-03 to 30-03 (period 2), 31-03 to 18-04 (period 3) and 19-04 to 12-06 (period 4).

**Figure A2:**
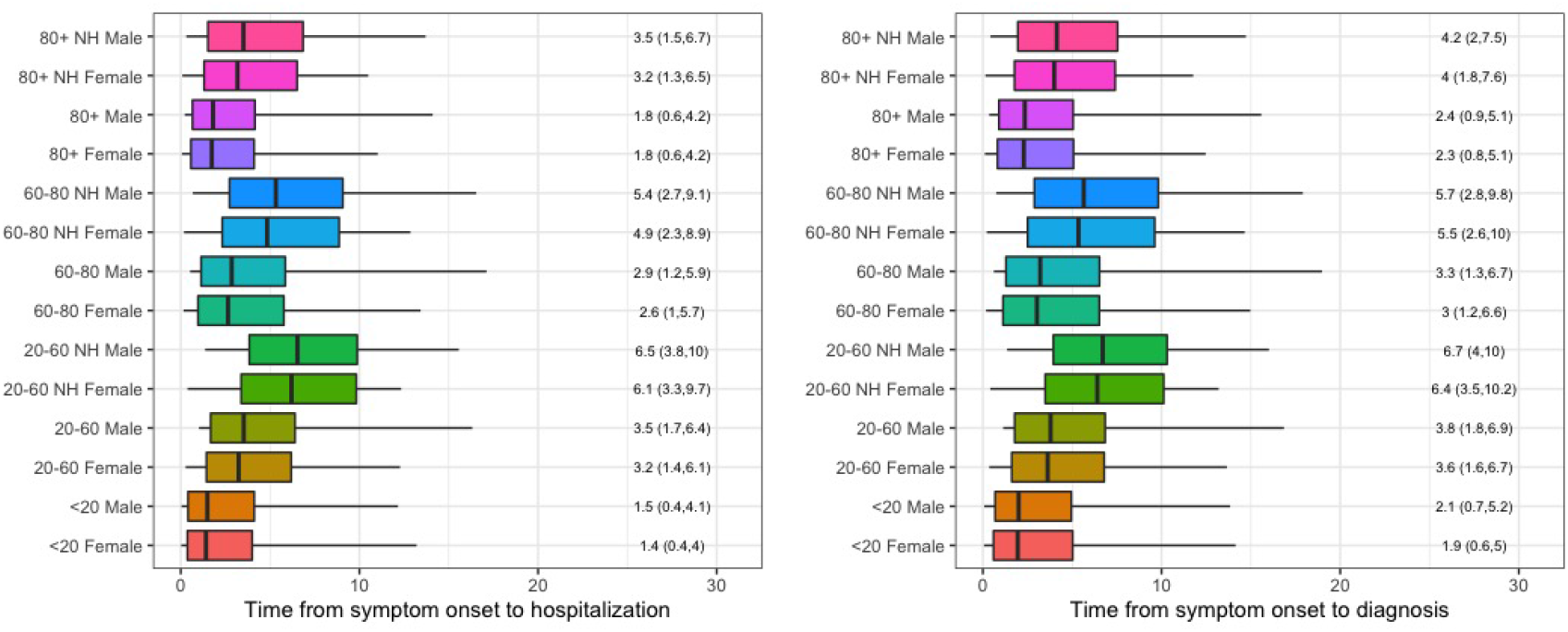
Comparison of delay distribution for different population groups. The boxplots show the estimated 5%, 25%, 50%, 75% and 95% quantiles, based on (untruncated) Weibull regression. Results are based on period 21-03 to 30-03.

**Table A1:** Summary of the reported delay time between symptom onset and hospitalization and between symptom onset and diagnosis

|  |  | Mean | Median | 2.5% | 25% | 75% | 97.5% | N |
| --- | --- | --- | --- | --- | --- | --- | --- | --- |
| Symptom onset - hospitalization |  |  |  |  |  |  |  |  |
| Overall |  | 5.6 | 5.0 | 0.0 | 2.0 | 8.0 | 20.0 | 12364 |
| Gender | Male | 5.9 | 5.0 | 0.0 | 2.0 | 8.0 | 20.0 | 6700 |
|  | Female | 5.2 | 4.0 | 0.0 | 1.0 | 7.0 | 20.0 | 5664 |
| Age | <20 | 3.0 | 1.0 | 0.0 | 0.0 | 4.0 | 14.0 | 227 |
|  | 20-60 | 6.8 | 7.0 | 0.0 | 3.0 | 9.0 | 20.0 | 4166 |
|  | 60-80 | 5.8 | 5.0 | 0.0 | 2.0 | 8.0 | 21.0 | 4717 |
|  | >80 | 3.9 | 2.0 | 0.0 | 0.0 | 6.0 | 16.0 | 3254 |
| Nursing home | Yes | 3.6 | 2.0 | 0.0 | 0.0 | 5.0 | 15.0 | 1863 |
|  | No | 5.9 | 5.0 | 0.0 | 2.0 | 8.0 | 20.0 | 5885 |
|  | Unknown | 5.9 | 5.0 | 0.0 | 2.0 | 8.0 | 21.0 | 4605 |
| Period | 01-03 to 20-03 | 4.7 | 4.0 | 0.0 | 1.0 | 7.0 | 14.0 | 1365 |
|  | 21-03 to 30-03 | 6.2 | 6.0 | 0.0 | 3.0 | 9.0 | 16.0 | 4593 |
|  | 31-03 to 18-04 | 5.6 | 4.0 | 0.0 | 1.0 | 8.0 | 21.0 | 4432 |
|  | 19-04 to 12-06 | 4.8 | 3.0 | 0.0 | 0.0 | 7.0 | 21.0 | 1974.0 |
| Symptom onset - diagnosis |  |  |  |  |  |  |  |  |
| Overall |  | 5.8 | 5.0 | 0.0 | 2.0 | 8.0 | 20.0 | 12574 |
| Gender | Male | 6.1 | 5.0 | 0.0 | 2.0 | 8.0 | 20.0 | 6798 |
|  | Female | 5.5 | 4.0 | 0.0 | 1.0 | 8.0 | 21.0 | 5776 |
| Age | <20 | 3.5 | 2.0 | 0.0 | 1.0 | 5.0 | 13.3 | 227 |
|  | 20-60 | 6.8 | 7.0 | 0.0 | 3.0 | 9.0 | 19.0 | 4131 |
|  | 60-80 | 6.1 | 5.0 | 0.0 | 2.0 | 8.0 | 21.0 | 4838 |
|  | >80 | 4.3 | 3.0 | 0.0 | 1.0 | 6.0 | 19.6 | 3378 |
| Nursing home | Yes | 3.6 | 2.0 | 0.0 | 1.0 | 5.0 | 15.0 | 1745 |
|  | No | 6.0 | 5.0 | 0.0 | 2.0 | 8.0 | 20.0 | 6190 |
|  | Unknown | 6.3 | 6.0 | 0.0 | 2.0 | 9.0 | 21.0 | 4628 |
| Period | 01-03 to 20-03 | 5.6 | 4.0 | 0.0 | 2.0 | 8.0 | 21.0 | 1629 |
|  | 21-03 to 30-03 | 6.6 | 7.0 | 0.0 | 3.0 | 9.0 | 17.0 | 4623 |
|  | 31-03 to 18-04 | 5.6 | 4.0 | 0.0 | 1.0 | 8.0 | 21.0 | 4427 |
|  | 19-04 to 12-06 | 4.4 | 3.0 | 0.0 | 0.0 | 7.0 | 21.0 | 1895 |

**Table A2:**
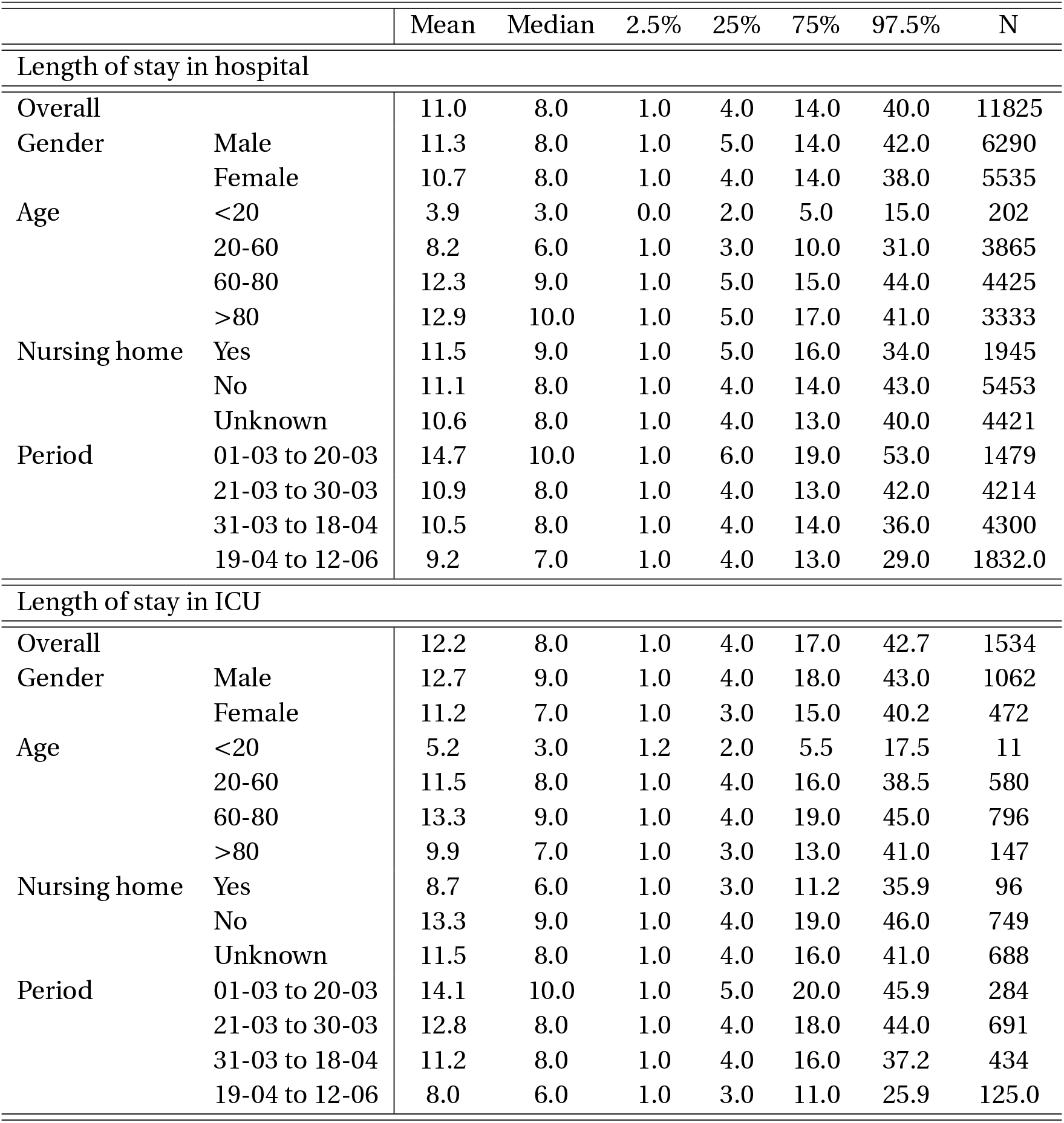
Summary of the reported length of stay in hospital

|  |  | Mean | Median | 2.5% | 25% | 75% | 97.5% | N |
| --- | --- | --- | --- | --- | --- | --- | --- | --- |
| Length of stay in hospital |  |  |  |  |  |  |  |  |
| Overall |  | 11.0 | 8.0 | 1.0 | 4.0 | 14.0 | 40.0 | 11825 |
| Gender | Male | 11.3 | 8.0 | 1.0 | 5.0 | 14.0 | 42.0 | 6290 |
|  | Female | 10.7 | 8.0 | 1.0 | 4.0 | 14.0 | 38.0 | 5535 |
| Age | <20 | 3.9 | 3.0 | 0.0 | 2.0 | 5.0 | 15.0 | 202 |
|  | 20-60 | 8.2 | 6.0 | 1.0 | 3.0 | 10.0 | 31.0 | 3865 |
|  | 60-80 | 12.3 | 9.0 | 1.0 | 5.0 | 15.0 | 44.0 | 4425 |
|  | >80 | 12.9 | 10.0 | 1.0 | 5.0 | 17.0 | 41.0 | 3333 |
| Nursing home | Yes | 11.5 | 9.0 | 1.0 | 5.0 | 16.0 | 34.0 | 1945 |
|  | No | 11.1 | 8.0 | 1.0 | 4.0 | 14.0 | 43.0 | 5453 |
|  | Unknown | 10.6 | 8.0 | 1.0 | 4.0 | 13.0 | 40.0 | 4421 |
| Period | 01-03 to 20-03 | 14.7 | 10.0 | 1.0 | 6.0 | 19.0 | 53.0 | 1479 |
|  | 21-03 to 30-03 | 10.9 | 8.0 | 1.0 | 4.0 | 13.0 | 42.0 | 4214 |
|  | 31-03 to 18-04 | 10.5 | 8.0 | 1.0 | 4.0 | 14.0 | 36.0 | 4300 |
|  | 19-04 to 12-06 | 9.2 | 7.0 | 1.0 | 4.0 | 13.0 | 29.0 | 1832.0 |
| Length of stay in ICU |  |  |  |  |  |  |  |  |
| Overall |  | 12.2 | 8.0 | 1.0 | 4.0 | 17.0 | 42.7 | 1534 |
| Gender | Male | 12.7 | 9.0 | 1.0 | 4.0 | 18.0 | 43.0 | 1062 |
|  | Female | 11.2 | 7.0 | 1.0 | 3.0 | 15.0 | 40.2 | 472 |
| Age | <20 | 5.2 | 3.0 | 1.2 | 2.0 | 5.5 | 17.5 | 11 |
|  | 20-60 | 11.5 | 8.0 | 1.0 | 4.0 | 16.0 | 38.5 | 580 |
|  | 60-80 | 13.3 | 9.0 | 1.0 | 4.0 | 19.0 | 45.0 | 796 |
|  | >80 | 9.9 | 7.0 | 1.0 | 3.0 | 13.0 | 41.0 | 147 |
| Nursing home | Yes | 8.7 | 6.0 | 1.0 | 3.0 | 11.2 | 35.9 | 96 |
|  | No | 13.3 | 9.0 | 1.0 | 4.0 | 19.0 | 46.0 | 749 |
|  | Unknown | 11.5 | 8.0 | 1.0 | 4.0 | 16.0 | 41.0 | 688 |
| Period | 01-03 to 20-03 | 14.1 | 10.0 | 1.0 | 5.0 | 20.0 | 45.9 | 284 |
|  | 21-03 to 30-03 | 12.8 | 8.0 | 1.0 | 4.0 | 18.0 | 44.0 | 691 |
|  | 31-03 to 18-04 | 11.2 | 8.0 | 1.0 | 4.0 | 16.0 | 37.2 | 434 |
|  | 19-04 to 12-06 | 8.0 | 6.0 | 1.0 | 3.0 | 11.0 | 25.9 | 125.0 |

**Figure A3:**
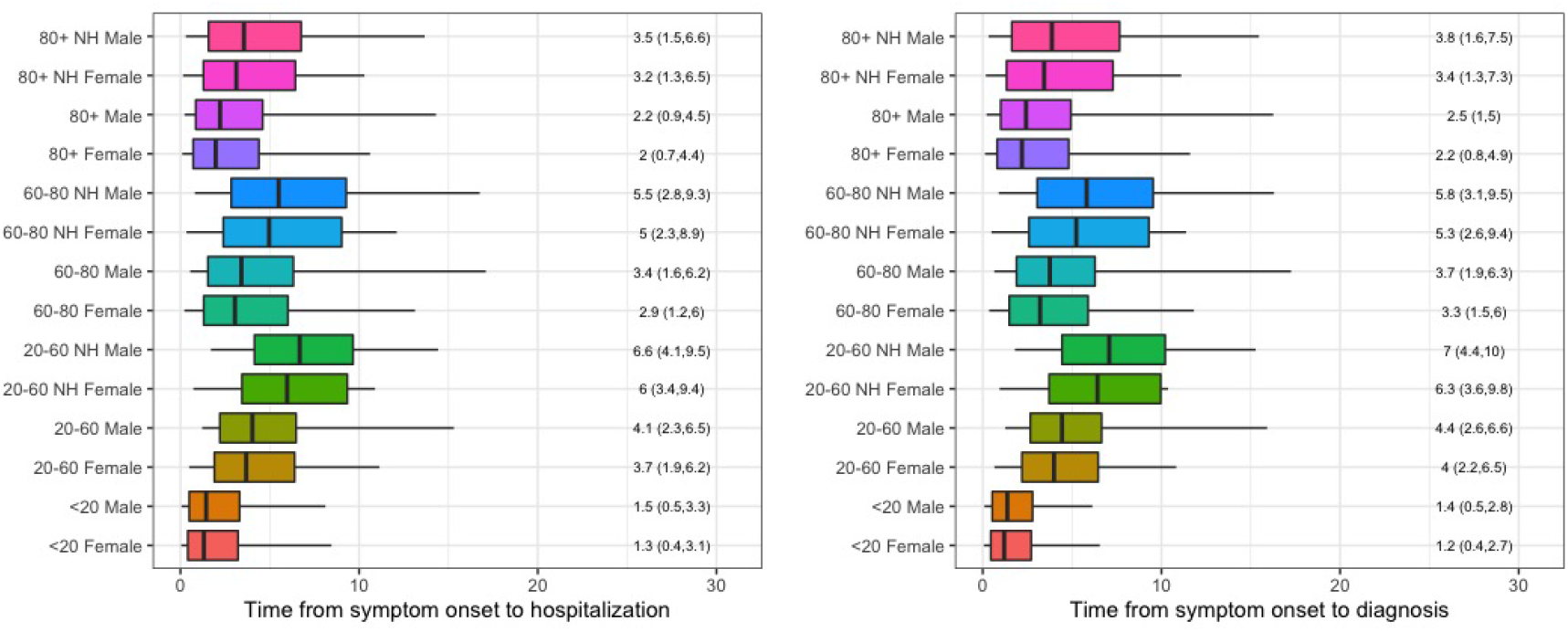
Comparison of delay distribution for different population groups. The boxplots show the estimated 5%, 25%, 50%, 75% and 95% quantiles, based on a truncated Weibull regression. The time intervals are assumed interval-censored in intervals (*x*_*i*_ − 1, *x*_*i*_ + 1). Results are based on period 21-03 to 30-03.

**Figure A4:**
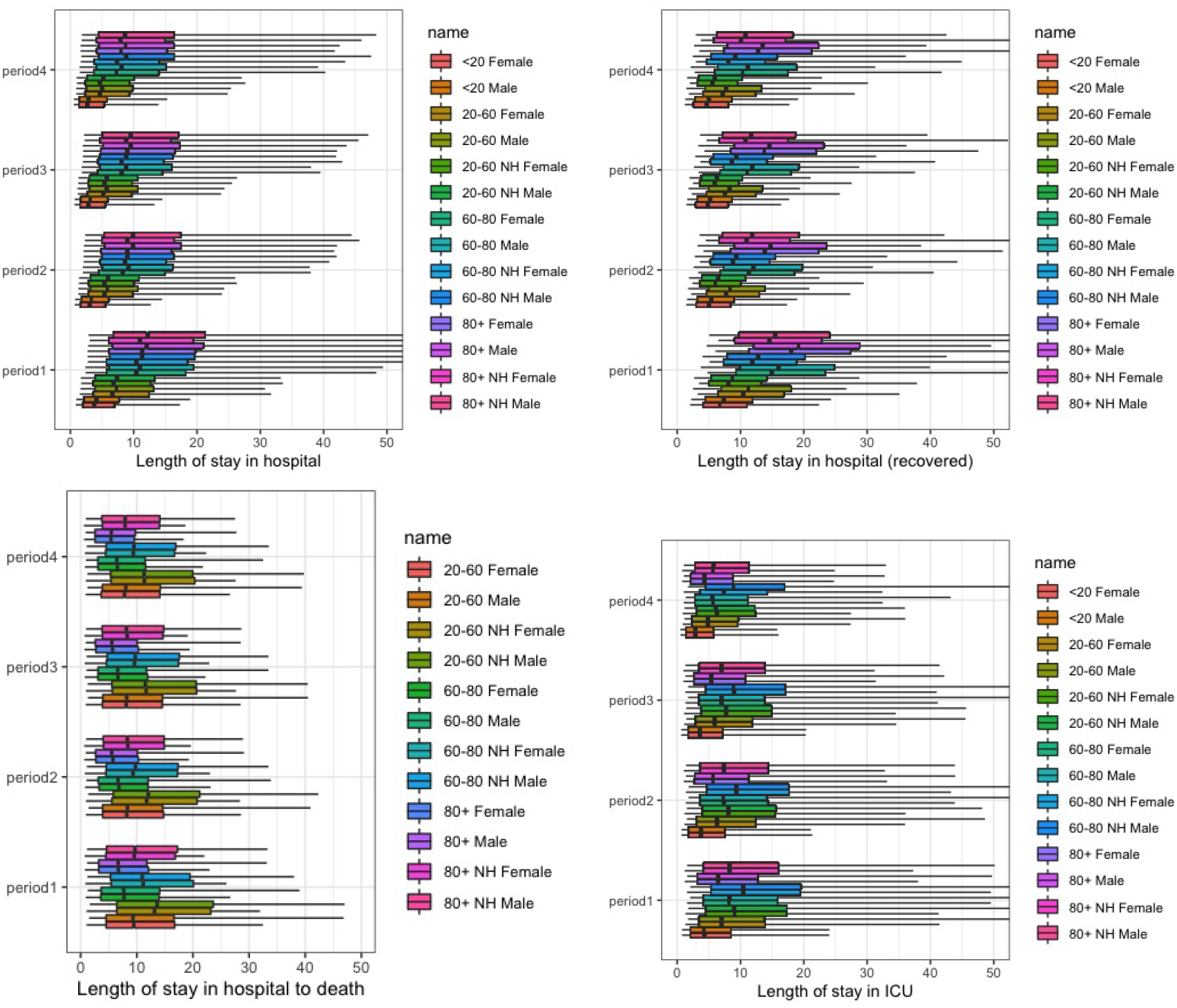
Comparison of delay distribution for different population groups. The boxplots show the estimated 5%, 25%, 50%, 75% and 95% quantiles, based on Weibull regression. Results are based on the time period 01-03 to 20-03 (period 1), 21-03 to 30-03 (period 2), 31-03 to 18-04 (period 3) and 19-04 to 12-06 (period 4).

**Figure A5:**
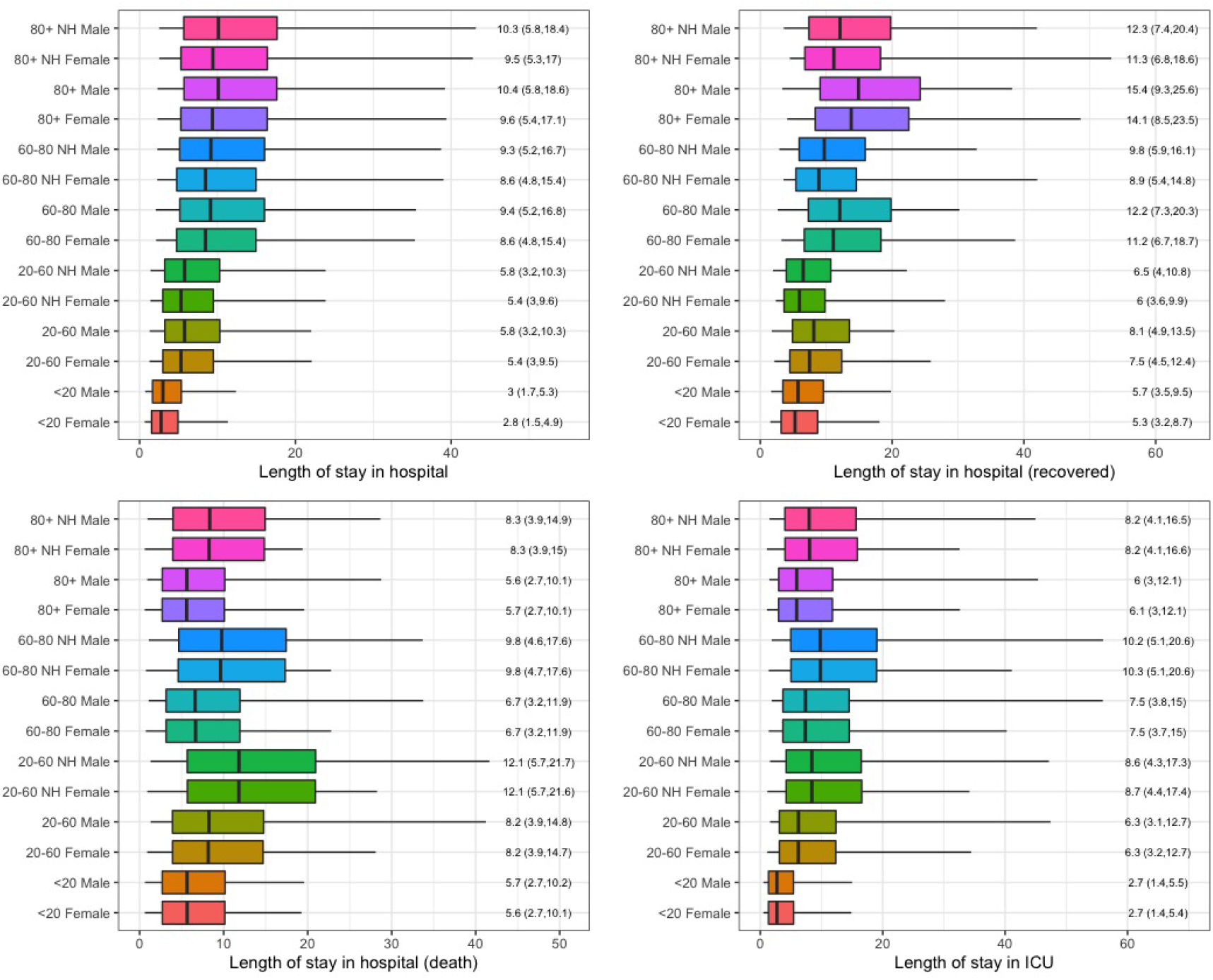
Comparison of delay distribution for different population groups. The boxplots show the estimated 5%, 25%, 50%, 75% and 95% quantiles, based on lognormal regression. The time intervals are assumed interval-censored in intervals (*x*_*i*_ − 1, *x*_*i*_ + 1). Results are based on period 21-03 to 30-03.

**Table A3:** List of members of the Belgian Collaborative Group on COVID-19 Hospital Surveillance and affiliations

| Name | Institution |
| --- | --- |
| Amir-Samy Aouachria | Centre Hospitalier Chrétien, Liège |
| Kristof Bafort | Mariaziekenhuis, Pelt |
| Leïla Belkhir | Cliniques Universitaires Saint-Luc, Brussels |
| Nathalie Bossuyt | Sciensano |
| Vincent Colombie | Centre Hospitalier Epicura, Baudour |
| Nicolas Dauby | Centre Hospitalier Universitaire Saint-Pierre, Brussels |
| Paul De Munter | Universitair Ziekenhuis Leuven, Leuven |
| Jessika Deblonde | Sciensano |
| Didier Delmarcelle | Clinique St. Jean, Brussels |
| Mélanie Delvallee | Centre Hospitalier de Wallonie Picarde, Tournai |
| Rémy Demeester | Centre Hospitalier Universitaire de Charleroi, Charleroi |
| Thierry Dugernier | Clinique Saint-Pierre, Ottignies |
| Xavier Holemans | Grand Hôpital de Charleroi, Charleroi |
| Frédéric Thys | Grand Hôpital de Charleroi, Charleroi |
| Benjamin Kerzmann | Clinique Notre Dame de Grâce, Gosselies |
| Pierre Yves Machurot | Centre Hospitalier de l'Ardenne |
| Philippe Minette | Centres Hospitaliers Jolimont |
| Jean-Marc Minon | Centre Hospitalier Régional de la Citadelle, Liège |
| Saphia Mokrane | Hôpitaux Iris Sud, Brussels |
| Catherine Nachtergal | Cliniques de l'Europe, Brussels |
| Séverine Noirhomme | Centre Hospitalier Régional de Namur |
| Denis Piérard | Universitair Ziekenhuis Brussel, Brussels |
| Camelia Rossi | Centre Hospitalier Universitaire Ambroise Paré, Mons |
| Carole Schirvel | CHIREC, Brussels |
| Erica Sermijn | A.S.Z. Ziekenhuis, Aalst |
| Frank Staelens | OLV Ziekenhuis, Aalst |
| Filip Triest | Algemeen Ziekenhuis Sint Lucas, Gent |
| Dominique Van Beckhoven | Sciensano |
| Nina Van Goethem | Sciensano |
| Jens Van Praet | Algemeen Ziekenhuis Sint Jan, Brugge-Oostende |
| Anke Vanhoenacker | Ziekenhuisnetwerk Antwerpen |
| Sarah Cooreman | Algemeen Ziekenhuis. Monica |
| Elise Willems | Algemeen Ziekenhuis Nikolaas, Sint-Niklaas |
| Chloé Wyndham-Thomas | Sciensano |

## Notes

### Competing Interest Statement

The authors have declared no competing interest.

### Funding Statement

H2020 Epipose (Epidemic intelligence to minimize 2019-nCoVs public health, economic and social impact in Europe)

### Author Declarations

UZ Gent (BC-07507)

